# Undetectable free aromatic amino acids in nails of breast carcinoma: Biomarkers discovery by a novel metabolite purification VTGE system

**DOI:** 10.1101/2019.12.22.19015669

**Authors:** Manmohan Mitruka, Charusheela R. Gore, Ajay Kumar, Sachin C Sarode, Nilesh Kumar Sharma

## Abstract

**BACKGROUND:** Metabolic reprogramming in breast cancer is depicted as a crucial change in the tumor microenvironment. Besides the molecular understanding of metabolic heterogeneity, appreciable attentions are drawn to characterize metabolite profiles in tumor tissue and derived biological fluids and tissue materials. Several findings reported on the metabolic alterations of free aromatic amino acids (FAAAs) and other metabolites in biological fluids. Furthermore, there is a significant gap in the development of a suitable method for the purification and analysis of metabolite biomarkers in nails of cancer patients.

**METHODS:** To address the metabolite alterations specifically FAAAs level in nails, fingernail clippings of breast cancer patients (N=10), and healthy subjects (N-12) were used for extraction and purification of metabolites. Here, we report a novel and specifically designed vertical tube gel electrophoresis (VTGE) system that helped for the purification of metabolites in the range of (100-1000 Da) from nail materials. The claimed VTGE system uses 15% polyacrylamide under non-denaturing and non-reducing conditions that makes eluted metabolites directly compatible with LC-HRMS and other analytical techniques. Qualitative and quantitative determination of FAAAs in nail lysates were done by positive ESI mode of Agilent LC-HRMS platform.

**RESULTS:** The analysis on collected data of nail metabolites clearly suggests that FAAAs including tryptophan, tyrosine, phenylalanine and histidine are undetectable in nail lysates of breast cancer over healthy subjects. This is a first report that shows the highly reduced levels of FAAAs in nails of breast cancer. Furthermore, present observation is in consonance with previous findings that support that breast cancer patients show cancer cachexia, high amino acid catabolism that drive metabolite driven cancer growth and proliferation.

**CONCLUSION:** This paper provides a proof of concept for a novel and specifically developed VTGE process that assisted to show the first evidence on undetectable level of FAAAs in nails of breast cancer patients as metabolite biomarkers. Here, the authors propose the potential use of VTGE assisted process to achieve metabolomic discovery in nails of breast cancer and other tumor types.

## INTRODUCTION

At global level, it is estimated that frequency of breast cancer among women is around 1.5 million each year. In the current decade, around 570,000 women died from breast cancer that is approximately 15% of all cancer deaths among women (1-2). Besides the need of better efficacious drug options with precision-guided therapy, a need for the early diagnosis and monitoring of cancer drug treatment is at the forefront at preclinical and clinical levels (3-4). To achieve the above goal, search for new classes of biomarkers at the levels of genome, transcriptome, small RNAs, proteome, and metabolome is highly relevant in a global context and Indian setting (5-6).

Intra- and inter-tumor heterogeneity at both cellular and non-cellular levels are depicted as the basis of origin, progression and drug response in breast cancer (7-10). The elucidation of metabolic heterogeneity at tumor microenvironment and macroenvironment level is important in the perspective of biomarkers development and future interventions in breast cancer (11-16). Among metabolic heterogeneity, amino acid catabolism that targets Free Aromatic Amino Acids (FAAAs) such as tryptophan, tyrosine, phenylalanine and histidine are regarded as one of the preferable hallmarks in breast cancer that support their growth and progression (17-22).

The determination of metabolite profiling is perceived as one viable option to detect early and late stage physiological and pathological changes in cancer patients (23-29). In this direction, several attempts are noticed by using metabolomic approaches, including LC-HRMS, GC-MS and NMR spectroscopy. These metabolomics approaches were focused on the tumor tissue and biological fluids, including serum, urine and saliva (30-36). However, literature completely lacks on the suitable methods that targets to profile nail metabolites of breast cancer and other tumor types.

In this paper, we claim a novel and specifically designed vertical tube gel electrophoresis (VTGE) based metabolite identifications of metabolites in nail lysates. Further, data report that undetectable levels of FAAAs in nail lysate of breast cancer patients may serve as potential diagnostic biomarkers.

## Methods

### Study Population

For this pilot study, breast cancer patients (N=10) and healthy women subjects (N=12) were recruited at Dr. D. Y. Patil Medical College, Pune, India. In accordance with the Institutional Ethics Committee (IEC), a formal approval was obtained to conduct research on healthy clinical subjects and breast cancer patients. All the participating clinical subjects were apprised about the aims of study and informed consent was collected.

In the present study, metabolite profiling data were analyzed between breast cancer patients and healthy subjects, without characterizing the molecular sub-types and prior chemotherapy. The primary goals were to develop a protocol for the preparation of the nail metabolite lysate, followed by purification process of nail metabolites and then finally characterizing with the help of the LC-HRMS. At this stage of study, we focused on the novel methods and processes that helped to collect metabolite profiles in the nail lysates of breast cancer and healthy subjects. In future, the claimed methods and processes that use VTGE and LC-HRMS technique may be used for large study population of breast cancer patients with attributes of molecular sub-types and history of treatments.

### Preparation of nail lysates

Fingernail clippings were collected in a microfuge tube and cleaned to remove debris, environmental contaminations by using a mild detergent and 70% ethyl alcohol. Further, fingernail clippings were dried, weighed and coded properly for healthy and breast cancer patients. Next, equal amount of fingernail clippings (20 mg) was added to 800 µl of extraction buffer (Tris-HCl (20mM, PH-8.5), 2.6M Thiourea, 5M Urea, Beta-Mercaptoethanol) for lysis of fingernail clippings. Next, sample mixture was incubated for 24 hr at 50□C under dark condition. Further, sample mixture was centrifuged at 15,000 X RPM for 30 min. The supernatant was collected in a fresh microfuge tube and filtered as by using 0.45 micron syringe filter membrane. The above prepared nail lysates of healthy and breast cancer patients were diluted three times, stored and labelled properly for purification by using VTGE system.

### Purification of metabolites by using VTGE system

In order to purify metabolites of nail materials, we employed a novel and specifically designed VTGE system that is standardized to use 15% polyacrylamide gel matrix to remove major components as proteins, polysaccharides, large lipid molecules and other various debris/contaminants. At the same time, claimed VTGE system allowed for purification of metabolites in the range of 100 Da to 1000 Da (37).

Above prepared sterile and filtered nail lysate (750 µl) was mixed with (250 µl) the loading buffer (4X Glycerol and Tris pH-6.8) and electrophoresed on VTGE casted with 15% acrylamide gel (acrylamide: bisacrylamide, 30:1) as a matrix. The fractionated elute was collected in electrophoresis running buffer that contains water and glycine and excludes traditional SDS, and other reducing agents. A notable distinctiveness of VTGE system claims that running buffer (3 ml) and elution buffer (3 ml) are identical as (Water-Glycine (192 mM), pH: 8.3). A flow diagram of VTGE method is presented in Figure 1A and 1B that show the assembly and design of VTGE system. Nail lysate metabolites from healthy and breast cancer patients were eluted in the same running buffer, which is referred as “Elution buffer” and placed in the lower electrophoretic tube that has anode wire. After loading of nail lysate (750 µl) along with loading buffer (4X, 250 µl), voltage and current ratio from power supply was maintained to generate 1500-2500 mW of power to achieve the electrophoresis of biological samples. The total run time was allowed for 2 hr. At the end of 2 hr, lower 3 ml collecting buffer was collected in a fresh microfuge tube for direct LC-HRMS characterization. Interestingly, pH of eluted metabolite buffer was measured for healthy and breast cancer patients that ranged between 3.0-3.5 acidic pH. An acidic pH (2.5-4.0) buffer containing metabolites are known to help in ionization efficiency during LC-HRMS analysis (38). At the end of the run, inner tube containing polyacrylamide gel was removed and placed for coomassie brilliant blue dye staining to ensure that protein components of nail lysates were trapped in the polyacrylamide matrix. Eluted nail metabolites from healthy and breast cancer patients were stored at -20°C for direct identification by LC-HRMS technique.

**Figure 1.**
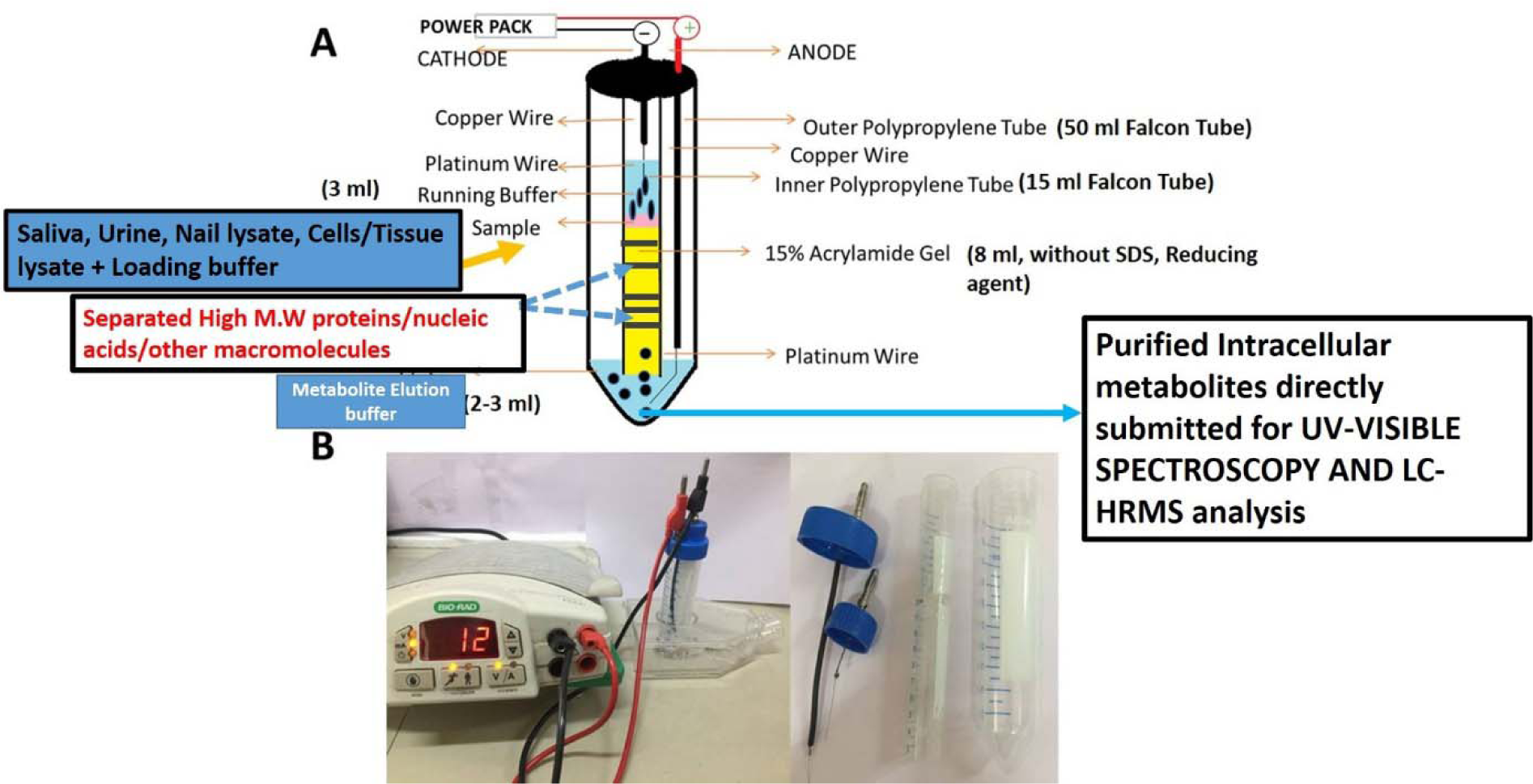
A flow diagram of novel and specifically designed vertical tube gel electrophoresis (VTGE) system for intracellular nail metabolite purification. Here, Figure 1A depicts the design, assembly and key features including nature of matrix, non-reducing and non-denaturing buffers. Figure 1B shows the working diagram that allow the nail metabolite purification from healthy subjects and breast cancer patients.

### Identification of potential nail metabolites by LC-HRMS

The qualitative and quantitative analysis of nail metabolites carried out by LC-HRMS. The purified nail metabolites from VTGE system was submitted to LC-HRMS. For liquid chromatography (LC) component, RPC18 column was used as Zorbax, 2.1 X 50 mm, 1.8 micron meters. Further, a flow rate of 0.2 ml/min and a gradient was formed by mixing mobile phase A (water containing 5mM ammonium acetate) and B (0.2% formic acid). For the run of sample, an injection volume was of 25 µl and a flow rate of solvent was maintained at 0.3 ml per minute. The HPLC column effluent was allowed to move onto an Electrospray Ionization Triple Quadrupole Mass Spectrometer (Agilent Technologies). For mass identification of nail metabolites, samples were run in positive electrospray ionization (ESI) M-H mode. During LC-HRMS analysis, mass spectrometer component was used as MS Q-TOF Quadrupole time-of-flight mass spectrometry (Q-TOF-MS) (Agilent Technologies, 6500 Series Q-TOF LC/MS System) with dual AJS electrospray ionization (ESI) mode. The acquisition mode of MS1 was recorded with a minimum value of m/z at 60 and maximum value of m/z at 1700.

## STATISTICAL ANALYSIS

Data are presented as the mean ± SD. The statistical significance between the healthy subjects and breast cancer patients were assessed with the help of one-way ANOVA test. Data calculation and statistical tests were performed by SPSS version 15.0 (SPSS) software package.

## RESULTS AND DISCUSSION

The importance of metabolite adaptations and profiling of metabolites in fine-needle aspiration biopsies, surgical biopsies, serum, urine and saliva was shown in various tumor types including breast, liver, thyroid and colorectal cancer (31-36). However, it is important to note that metabolite profiling in nail materials of cancer patients, including breast cancer has not been reported in literature. Besides non-availability of nail metabolite profiling in cancer patients, even a suitable method and processes to profile metabolites from a nail lysate of other human disease is completely lacking.

### Novelty of VTGE process

In literature, limited attempts are available that delineate the nail metabolite profiling in an environmental exposure cases and disease conditions (39-44). Interestingly, a metabolomic study of cancer metabolite profiles in nail lysate has not been investigated previously in the literature. To achieve a new and additional knowledge on metabolomic biomarkers in breast cancer, this paper claims a novel and specifically designed VTGE system and involved methods to purify nail lysate metabolites. The lysates are directly compatible with LC-HRMS techniques without any further need of complicated extraction, modifications and labelling protocols to identify and estimate metabolite biomarkers. Among a pool of various metabolites, FAAAs are considered as key tumor metabolite that supports growth and progression (17-22, 30). Based on the literature, there is not a single paper that addresses the importance of FAAAs as potential biomarkers in nail materials of breast cancer patients. To address the need of non-invasive, cheaper and rapid methods for the metabolite biomarkers in breast cancer, the authors present a novel and specifically designed VTGE based purification of metabolites (100-1000 Da) of nail materials of breast cancer patients. The claimed VTGE method is simply the out-of-box innovative applications with respect to classical Laemmli (1970)^45^ system that is primarily used for the separation and purification of large macromolecules such as proteins and nucleic acids. In this novel method, the authors have designed a VTGE system with the help of laboratory plastic ware that purifies nail metabolites (100 Da-1000 Da) by direct elution in the lower running buffer.

### Nails accumulate metabolites

Recently, metabolomic studies in various biological fluids, including serum, urine, saliva and tissues are reported to highlight the importance of metabolite as biomarkers (15, 16; 23-29). However, use of nails as a source of metabolite biomarkers in cancer and other human disease conditions are highly limited. On the other hand, use of nail metabolite profiling is reported to some extent in case of toxicology and environmental exposure. Limited reports support the avenues to explore nail metabolites, including ethyl glucuronide in the keratinous matrices of nail materials of patients (40-44). It is worth to mention that glucuronide derivatives as a hallmarks of liver metabolism upon drugs. Therefore, presence of ibuprofen glucuronide in nails of healthy and breast cancer patients is supported by the previous reports on detection of ethyl glucuronide in the keratinous matrices of nails. In other way, claimed VTGE system assisted nail metabolite profiling approach is convincing and in line with existing, but highly limited reports. Furthermore, acid digestion and SPE clean-up of metabolite derived from alternative plasticizers and flame retardants such as bis(2-propylheptyl) phthalate (DPHP), bis(2-ethylhexyl) terephthalate (DEHTP), bis(2-ethylhexyl) adipate (DEHA)), and one FR (2,2-bis (chloromethyl)-propane-1,3-diyltetrakis(2-chloroethyl) bisphosphate (V6) in nail materials of selected participants (40,42). A nail metabolite profiling study reported the use of ball mill grounding and extraction process followed by UHPLC-triple quadrupole-mass analyzer (41).

In the light of negligible approaches to profile nail metabolites in breast cancer and other disease conditions, we claim that VTGE system assisted purification process is novel and highly efficient with LC-HRMS technique compared to scarce metabolite identification approaches from nails. Another curious fact is important to be discussed that the use of nails as a source of metabolite biomarkers in breast cancer patients and other pathological conditions may have an added advantage over other biological fluids such as serum, saliva and urine in terms of less microbial and environmental contaminations. The data clearly show that identified nail metabolites, including FAAAs and drug metabolites are detected in positive ESI total ion chromatogram of healthy subjects (Figure 2A) that show Tryptophan (RT-1.523), L-Phenylalanine (RT-5.76), L-Tyrosine (RT-2.198), Histidine (RT-0.624). On the other hand, positive ESI total ion chromatogram of breast cancer patients (Figure 2B) indicates on presence of anti-cancer drug metabolites namely 6a,3’-p-Dihydroxypaclitaxel (RT-8.493) and Doxorubicinol (RT-10.026). In fact, ibuprofen glucuronide (RT-8.607), a well-known metabolite of anti-inflammatory drug ibuprofen is detected in both healthy subjects and breast cancer patients that clearly convince the precision and efficacy to show the proof-of-concept of VTGE assisted nail metabolites purification and identifications by LC-HRMS. Finally, positive ESI chromatogram reveals undetectable FAAAs metabolites in nails of breast cancer patients compared to highly abundant in the case of healthy subjects.

**Figure. 2.**
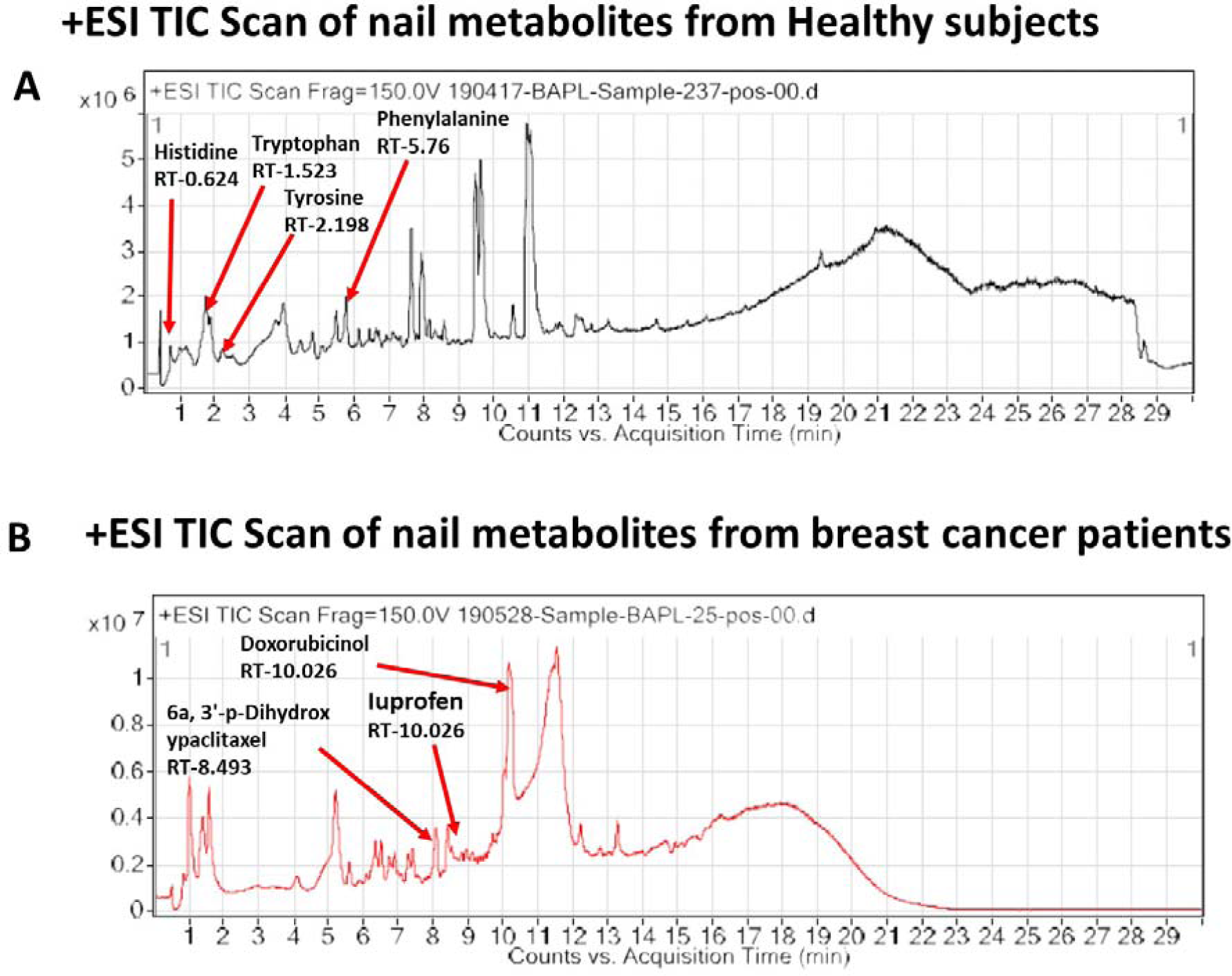
VTGE purified nail metabolites identified by LC-HRMS show unique TIC in healthy subjects over breast cancer patients To identify and profile nail metabolites changes, VTGE system assisted purified nail metabolites were directly submitted to LC-HRMS with positive ESI mode. Total ion chromatogram (TIC) of purified nail lysate of healthy subjects breast cancer patients is given in Figure 2A and Figure 2B, respectively.

### Identifications of drug metabolites in nail

Furthermore, we looked into the LC-HRMS identified metabolites other than FAAAs and the data suggest that various kinds of drugs including anti-inflammatory, anti-analgesic, antibiotics and anti-cancer drugs are actually accumulated in the nails of selected clinical subjects. Some of notable anticancer drug metabolites such as 6a, 3’-p-Dihydroxypaclitaxel and doxorubicinol are exclusively highly abundant in nail lysates of most of selected cancer patients. A positive ESI extracted ionization chromatogram (EIC) is presented as 6a, 3’-p-Dihydroxypaclitaxel (+ESI EIC 871.2682, 872.2716, 889.2788, 890.2821) and doxorubicinol(+ESI EIC 527.1786, 528.1819, 545.1892, 546.1925) in Figure 3B and Figure 3C, respectively. Furthermore, a positive ESI EIC of ibuprofen glucuronide (+ESI EIC 364.1517, 382.1622), a well-known anti-inflammatory drug is identified in both healthy subjects and breast cancer patients (Figure 3A, Table 1B). Further, abundance, mass value, m/z value and other parameters of identified drugs metabolites in nails of selected healthy and breast cancer patients is given in Table 1 and 2. On the other side, anti-inflammatory drug metabolites such as ibuprofen glucuronide is detected in both healthy and cancer patients and that strongly support the precision and reproducibility of claimed VTGE process for the improved precision and efficacy during LC-HRMS metabolite identifications. Besides above identified FAAAs and drug metabolites, VTGE assisted purified nail metabolite reveals the presence of smallest size metabolite 3-methylbut-3-enoic acid with a m/z value of 101.6 and a mass value (100.0524 Da). On the other hand, the highest mass metabolite is detected in nails as 28-Glucosylarjunolate 3-[rhamnosyl-(1->3)-glucuronide] with m/z a 973.4944 and a mass value (973.4944 Da). Here, the authors would like to draw attention that these two metabolites are known to be secreted in urine. Therefore, our claim on using VTGE assisted nail metabolite purifications is further strengthened. Interestingly, mass range of identified nail metabolites matches with the claim VTGE assisted purification approaches and that encourages to explore the distinct nail metabolite profiling between healthy and breast cancer patients. Here, the authors point out that detection of these unique metabolized products and drug metabolites in the nail lysates of healthy and breast cancer patients is a first report in the literature. To bring the evidence on the presence of drugs and their metabolites corroborate the chance of success with reference to the highly distinct abundance of FAAAs in healthy subjects over breast cancer patients.

**Table 1A.** List of free aromatic amino acids and their abundance in nail lysates.

| Sr. No | Name of Free aromatic amino acids | Healthy subjects* | Breast cancer patients |
| --- | --- | --- | --- |
| 1. | L/D-Tryptophan | 18407 ± 5071 | Undetectable |
| 2. | L-Phenylalanine | 3012017 ± 80100 | Undetectable |
| 3. | L-Tyrosine | 193292 ± 40817 | Undetectable |
| 4. | Histidine | 46544 ± 8981 | Undetectable |

\* Data represent mean ± S.D (N=10 Breast cancer patients and N=12 for healthy subjects)

**Table 1B.**
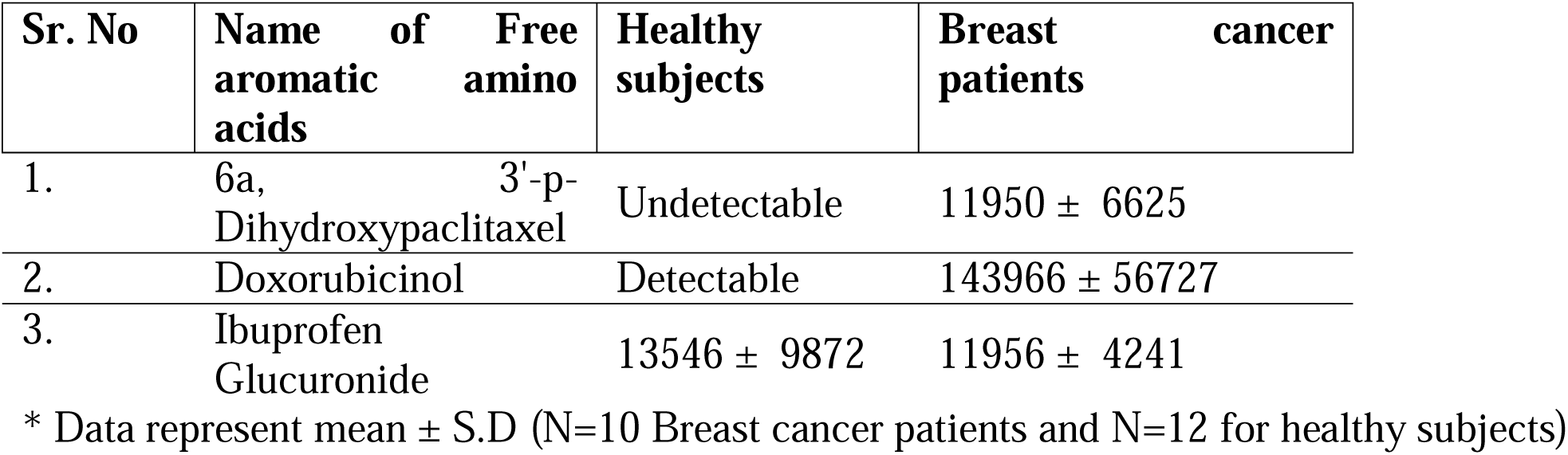
List of drugs and derived metabolites and their abundance in nail lysates.

**Table 2.**
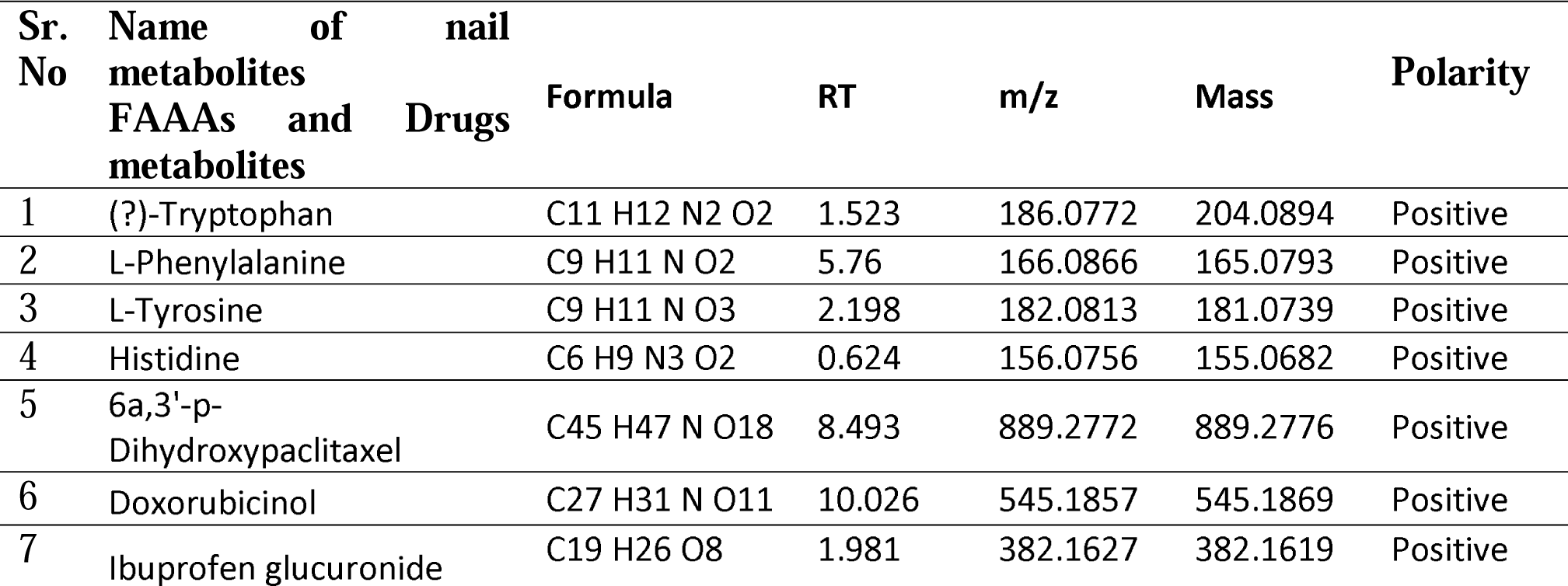
List of free aromatic amino acids and their abundance in nail lysates.

**Figure. 3.**
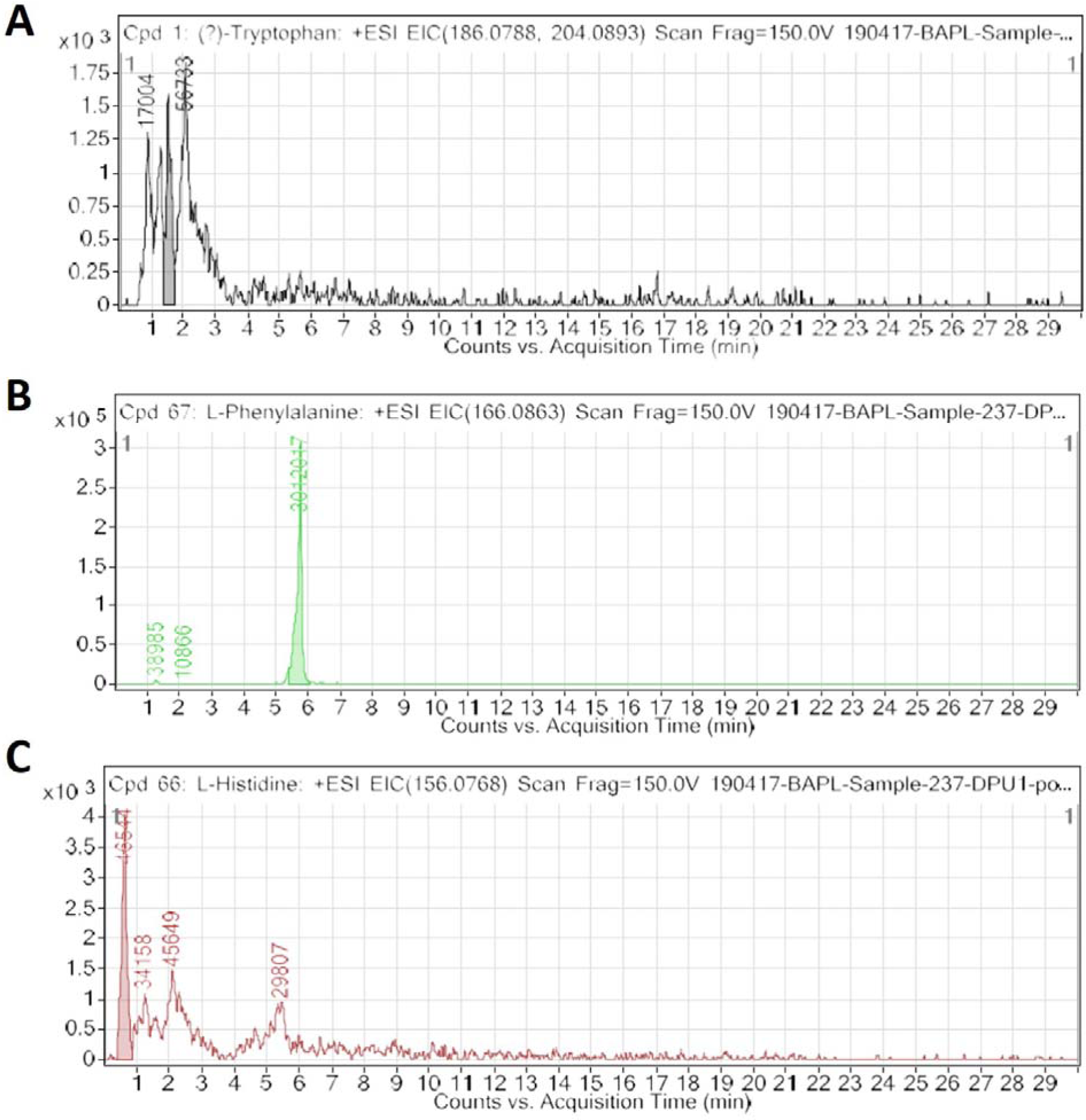
An extracted ion chromatogram (EIC) of drugs metabolized products in nail lysates, a first report is distinctively detected in healthy subjects and breast cancer patients. Here, Figure 3A, Figure 3B and Figure 3C represent a +ESI EIC of ibuprofen glucuronide (+ESI EIC 364.1517, 382.1622), 6a, 3’-p-Dihydroxypaclitaxel (+ESI EIC 871.2682, 872.2716, 889.2788, 890.2821) and doxorubicinol (+ESI EIC 527.1786, 528.1819, 545.1892, 546.1925), respectively.

### Reduction in level of FAAAs in breast cancer

Among the key role of amino acid metabolism in tumor microenvironment, a role of tryptophan and other aromatic amino acid catabolism is suggested that support immunosuppressive landscape (17-22; 30-32). In essence, amino acid catabolism including of tryptophan plays an important role in immune heterogeneity in tumor niche. The metabolized product of tryptophan as kynurenine is suggested to promote differentiation of T regulatory cells that help in immunosuppression. At the molecular level, one of two enzymes indoleamine 2,3-dioxygenase (IDO-1/-2) or tryptophan 2,3-dioxygenase 2 (TDO2) is known to initiate catabolism of tryptophan that lead to the generation of l-kynurenine (21,22,36). In the present paper, we show the clear and novel evidence on the appreciable level of FAAAs including tryptophan, phenylalanine, tyrosine and histidine in the nail lysate of healthy subjects (Table 1A). Interestingly, among FAAAs, phenylalanine is found to be highest in abundance,then followed by tyrosine, histidine and tryptophan. On the other hand, level of FAAAs is undetectable in the nail lysate of breast cancer patients. Here, the claimed methods and process is suggested to reveal undetectable levels of FAAAs in the nail lysate of breast cancer patients as potential metabolite biomarkers over healthy subjects (Figure 4, Table 1 and Table 2). A representative positive EIC of tryptophan (+ESI EIC 186.0788, 204.0893), phenylalanine (+ESI EIC 166.0863) and histidine (+ESI EIC 156.0768) is illustrated in Figure 4A, Figure 4B and Figure 4C, respectively. Based on the TIC and EIC of identified FAAAs in the nails of healthy subjects and at the same time undetectable in breast cancer patients is vividly determined quantitatively by LC-HRMS techniques. The highly efficient mass ionization of FAAAs identified in nails is credited to the novel purification methods assisted by VTGE system. Indeed, this observation is in consonance with the existing views on altered amino acid catabolism and their derived products support tumor growth and progression.

**Figure. 4.**
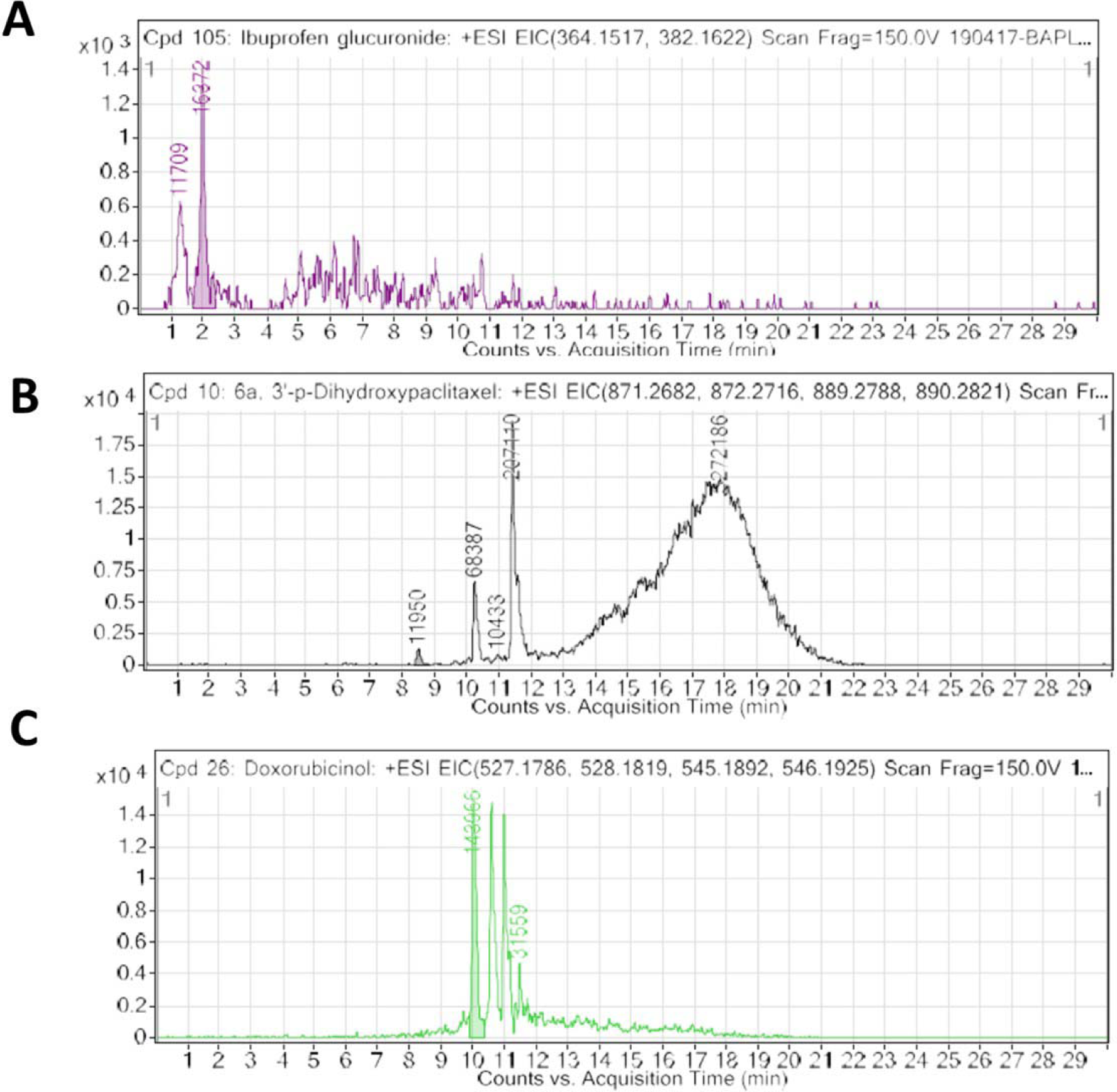
An extracted ion chromatogram (EIC) of free aromatic amino acids (FAAAs) in nail lysates, a novel report that shows in highly abundant in healthy subjects and undetectable in breast cancer patients. Here, Figure 4A, Figure 4B and Figure 4C represent a +ESI EIC of Tryptophan (+ESI EIC 186.0788, 204.0893), Phenylalanine (+ESI EIC 166.0863) and histidine (+ESI EIC 156.0768), respectively.

**Figure 5.**
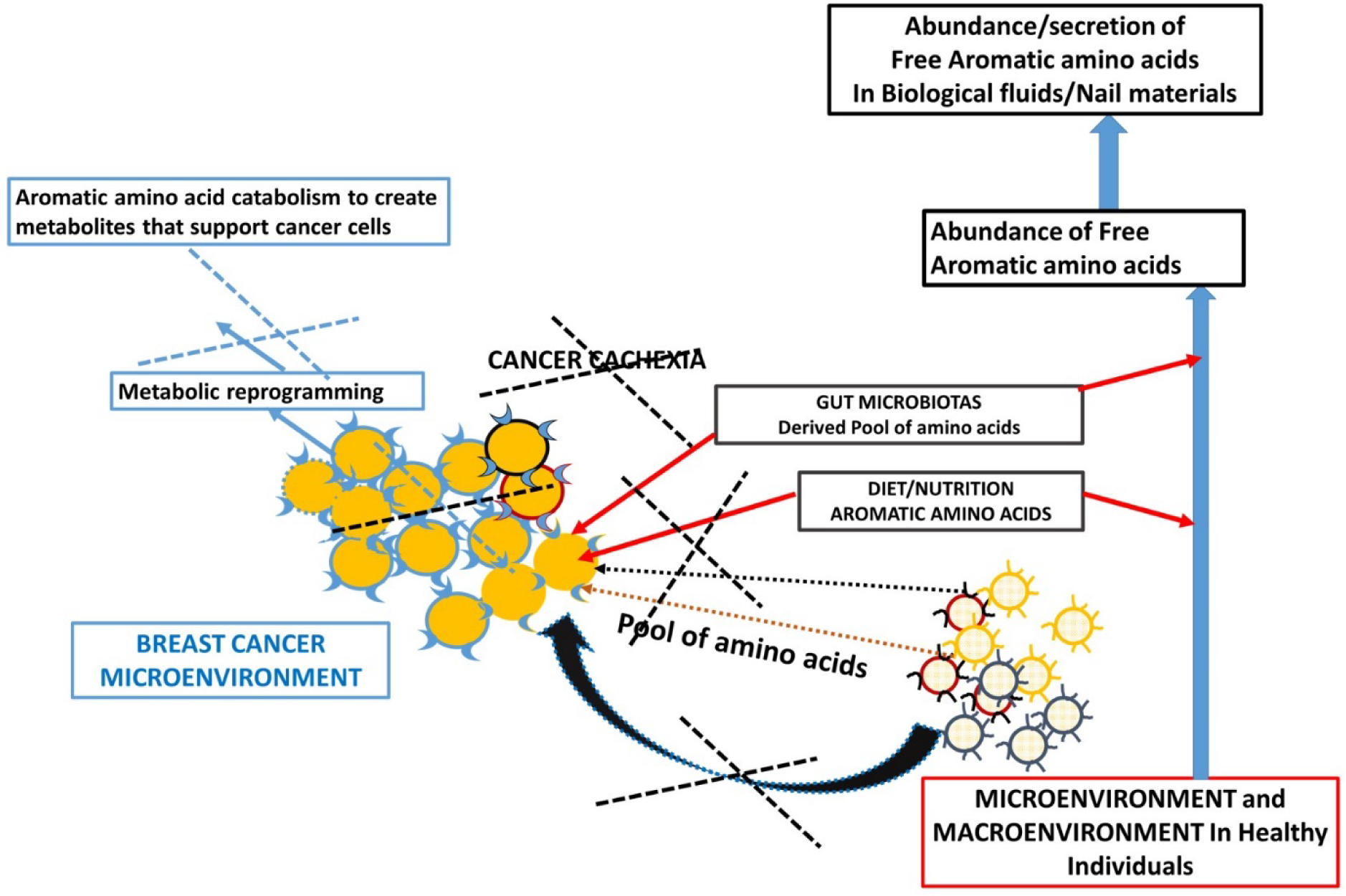
A proposed model of understanding that support the high abundance of FAAAs in biological fluids and tissue materials including nails of healthy subjects.

**Figure 6.**
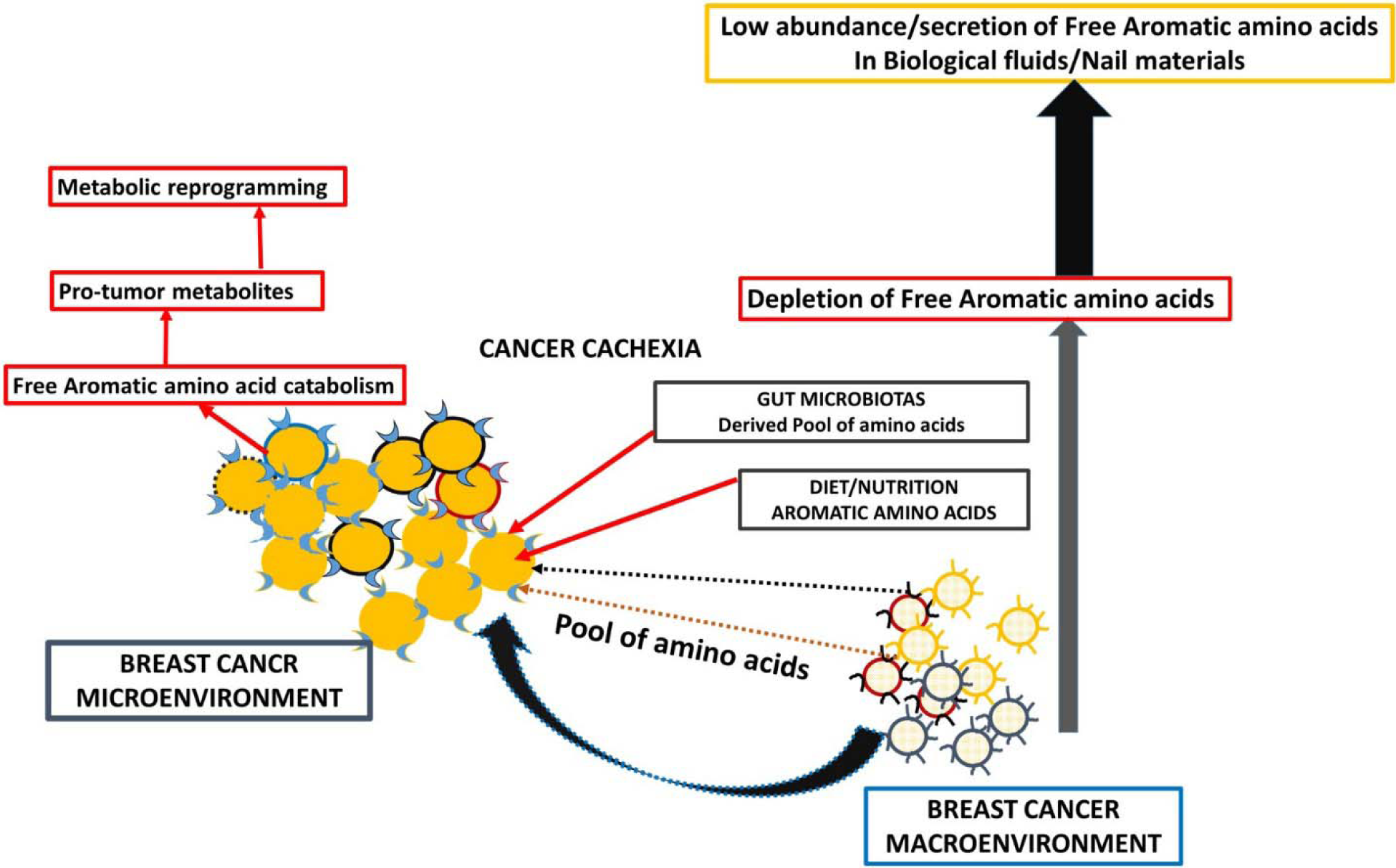
A proposed model of metabolic reprogramming in breast cancer microenvironment and macroenvironment that leads to FAAAs catabolism and in turn undetectable level of FAAAs in nails and other non-tumor biological fluids and materials.

Metabolic dependency and addiction to use the amino acid by cancer cells are known to support self-sustained growth and aggressiveness (23-29). In case of breast cancer patients, abundance of key amino acids, including FAAA in the other body parts and biological fluids is reduced. Furthermore, this metabolic adaptation in breast cancer patients may be a factor behind the undetectable levels of FAAAs compared to healthy subjects. At the same time, availability of FAAA in the tumor mass is elevated and also the catabolized products of these amino acids are increased (6,26-29). A metabolomic study in pancreatic and ovarian cancer indicates the glutamine citrulline, and histidine can serve as a potential biomarker (25,29). In prostate, colorectal and breast cancers level of neopterin and tryptophan metabolized metabolites in serum is suggested as predictive and prognostic factors (21,30).

The reduction of FAAAs in the nail lysate of breast cancer patients compared to healthy subjects is well connected with a recent study that points out the low level of serum free amino acids, including arginine, alanine, isoleucine, tyrosine and tryptophan in breast cancer (stage I-III) patients (33). A study by liquid chromatography-tandem mass spectrometry (LC-MS/MS) metabolic profiling and bioinformatics analysis, alterations in arginine/proline metabolism, tryptophan metabolism, and fatty acid biosynthesis are observed in breast cancer patients (26).

A metabolite profiling suggests that tyrosine, phenylalanine, or tryptophan decrease in the serum of patients with gastroesophageal cancer. At the same time, level of FAAAs such as tyrosine, phenylalanine and tryptophan is higher in tumor tissue of gastroesophageal cancer (20). The bioavailability of plasma free amino acids is shown to be implicated in alerting the tumor protein synthesis that is widely considered as one of tumor hallmarks. In fact, the profile of plasma free amino acids, especially, aromatic amino acids tyrosine, phenylalanine, tryptophan and histidine is suggested to be changed according to the stages and types of cancer. In another way, intra- and inter tumor heterogeneity may play a role in the distinct profile of free aromatic amino acids (18). Furthermore, a metabolic study supported by NMR spectroscopy indicates the occurrence of elevated glycolytic pathway and low abundance of glutamine in the plasma samples of brain tumors (24). In summary, existing views on amino acid catabolism as a hallmark of breast and other cancer types is supported by an additional and crucial evidence that support that breast cancer patients have undetected levels of FAAAs in nails compared to healthy subjects. Wherein, the accumulation of FAAAs in nails of healthy subjects should be viewed in line with the metabolic landscape during normal physiological conditions and well supported by selected metabolic study in other biological fluids.

## CONCLUSION

In spite of emanating understanding on genetic, epigenetic and environmental factors that impact tumorigenesis, a role of metabolic reprogramming is highly appreciated in tumor heterogeneity. A better understanding about metabolic reprogramming is crucial for prognostic markers, diagnostic avenue, therapeutic monitoring and combinatorial drug therapy. In this direction, use of a metabolomic approach to characterize metabolites qualitatively and quantitatively in various tissue samples and non-invasive biological fluids/materials including nails is emphasized at preclinical and clinical levels. Based on the above needs and available avenues, claimed methods that use VTGE system based purification of nail metabolites and their precise estimation by LC-HRMS promote as a novel and the first report on breast cancer metabolomic study. Furthermore, the described methods and process is warranted to facilitate metabolite biomarker study in other biological fluids/tissues in various cancer models. Hence, distinct level of FAAAs in nails of healthy subjects to breast cancer patients is warranted to be developed as a clinical assay in the future.

## Data Availability

Data will be made available upon request.

## ACKNOWLEDGEMENTS

The authors acknowledge financial support from DST-SERB, Government of India, New Delhi,India (SERB/LS-1028/2013) and Dr. D.Y. Patil Vidyapeeth, Pune, India (DPU/05/01/2016).

## Notes

### Competing Interest Statement

The authors have declared no competing interest.

### Funding Statement

This work is funded by DPU, Pune, India

## REFERENCE

1. Bray F, Ferlay J, Soerjomataram I, Siegel RL, Torre LA, Jemal A. Global cancer statistics 2018: GLOBOCAN estimates of incidence and mortality worldwide for 36 cancers in 185 countries. CA Cancer J Clin. 2018. 68(6):394–424.

2. GBD 2017, Disease and Injury Incidence and Prevalence Collaborators. Global, regional, and national incidence, prevalence, and years lived with disability for 354 diseases and injuries for 195 countries and territories, 1990-2017: a systematic analysis for the Global Burden of Disease Study 2017. Lancet. 2018. 392(10159):1789–1858.

3. Ganapathy V, Thangaraju M, Prasad PD. Nutrient transporters in cancer: relevance to Warburg hypothesis and beyond. Pharmacol Ther 2009. 121:29–40.

4. Hanahan D, Weinberg RA. Hallmarks of cancer: the next generation. Cell. 2011. 144(5):646–74.

5. Pavlova NN, Thompson CB. The Emerging Hallmarks of Cancer Metabolism. Cell Metab. 2016. 23(1):27–47.

6. Lukey MJ, Katt WP, Cerione RA. Targeting amino acid metabolism for cancer therapy. Drug Discov Today. 2017. 22(5):796–804.

7. Nilendu P, Sarode SC, Jahagirdar D, Tandon I, Patil S, Sarode GS. et al. Mutual concessions and compromises between stromal cells and cancer cells: driving tumor development and drug resistance. Cellular Oncology. 2018. 41(4):353–367.

8. Pan Y, Cao M, Liu J, Yang Q, Miao X, Go VLW. et al. Metabolic Regulation in Mitochondria and Drug Resistance. Adv Exp Med Biol. 2017. 1038:149–171.

9. Patel H, Nilendu P, Jahagirdar D, Pal JK, Sharma NK. Modulating non-cellular components of microenvironmental heterogeneity: A masterstroke in tumor therapeutics. Cancer Biology & Therapy. 2018. 19(1):3–12.

10. Yachida S, Mizutani S, Shiroma H, Shiba S, Nakajima T, Sakamoto T. et al. Metagenomic and metabolomic analyses reveal distinct stage-specific phenotypes of the gut microbiota in colorectal cancer. Nat Med. 2019. 25(6):968–976.

11. Wu H, Xue R, Lu C, Deng C, Liu T, Zeng H, et al. Metabolomic study for diagnostic model of oesophageal cancer using gas chromatography/mass spectrometry. J Chromatogr B 2009. 877:3111–7.

12. Cheng Y, Xie G, Chen T, Qiu Y, Zou X, Zheng M. et al. Distinct urinary metabolic profile of human colorectal cancer. J Proteome Res. 2012. 11(2):1354–63.

13. Rodrigues D, Jerónimo C, Henrique R, Belo L, de Lourdes Bastos M, de Pinho PG, Carvalho M. Biomarkers in bladder cancer: A metabolomic approach using in vitro and ex vivo model systems. Int J Cancer. 2016. 139(2):256–68.

14. Hart CD, Tenori L, Luchinat C, Di Leo A. Metabolomics in Breast Cancer: Current Status and Perspectives. Adv Exp Med Biol. 2016. 882:217–34.

15. Lario S, Ramírez-Lázaro MJ, Sanjuan-Herráez D, Brunet-Vega A, Pericay C, Gombau L, Junquera F, Quintás G, Calvet X. Plasma sample based analysis of gastric cancer progression using targeted metabolomics. Sci Rep. 2017. 7(1):17774.

16. Kaushik AK, DeBerardinis RJ. Applications of metabolomics to study cancer metabolism. Biochim Biophys Acta Rev Cancer. 2018. 1870(1):2–14.

17. Bozzetti F, Migliavacca S, Scotti A, Bonalumi MG, Scarpa D, Baticci F, et al. Impact of cancer, type, site, stage and treatment on the nutritional status of patients. Ann Surg 1982. 196:170–9.

18. Lai H-S, Lee J-C, Lee P-H, Wang S-T, Chen W-J. Plasma free amino acid profile in cancer patients. Semin Cancer Biol 2005. 15:267–76.

19. Nagata C, Wada K, Tsuji M, Hayashi M, Takeda N, Yasuda K. Plasma amino acid profiles are associated with biomarkers of breast cancer risk in premenopausal Japanese women. Cancer Causes Control. 2014. 25(2):143–9.

20. Wiggins T, Kumar S, Markar SR, Antonowicz S, Hanna GB. Tyrosine, phenylalanine, and tryptophan in gastroesophageal malignancy: a systematic review. Cancer Epidemiol Biomarkers Prev. 2015. 24(1):32–8.

21. Puccetti P, Fallarino F, Italiano A, Soubeyran I, MacGrogan G, Debled M. et al. Accumulation of an endogenous tryptophan-derived metabolite in colorectal and breast cancers. PLoS One. 2015. 10(4):e0122046.

22. Amobi A, Qian F, Lugade AA, Odunsi K. Tryptophan Catabolism and Cancer Immunotherapy Targeting IDO Mediated Immune Suppression. Adv Exp Med Biol. 2017. 1036:129–144.

23. Jee SH, Kim M, Kim M, Yoo HJ, Kim H, Jung KJ, Hong S, Lee JH. Metabolomics Profiles of Hepatocellular Carcinoma in a Korean Prospective Cohort: The Korean Cancer Prevention Study-II. Cancer Prev Res (Phila). 2018. 11(5):303–312.

24. Baranovičová E, Galanda T, Galanda M, Hatok J, Kolarovszki B, Richterová R, Račay P. Metabolomic profiling of blood plasma in patients with primary brain tumours: Basal plasma metabolites correlated with tumour grade and plasma biomarker analysis predicts feasibility of the successful statistical discrimination from healthy subjects - a preliminary study. IUBMB Life. 2019. 71(12):1994–2002.

25. Fest J, Vijfhuizen LS, Goeman JJ, Veth O, Joensuu A, Perola M, Männistö S. et al. Search for Early Pancreatic Cancer Blood Biomarkers in Five European Prospective Population Biobanks Using Metabolomics. Endocrinology. 2019. 160(7):1731–1742.

26. Jasbi P, Wang D, Cheng SL, Fei Q, Cui JY, Liu L. Breast cancer detection using targeted plasma metabolomics. J Chromatogr B Analyt Technol Biomed Life Sci. 2019. 1105:26–37.

27. Lécuyer L, Dalle C, Lyan B, Demidem A, Rossary A, Vasson MP, Petera M. et al. Plasma Metabolomic Signatures Associated with Long-term Breast Cancer Risk in the SU.VI.MAX Prospective Cohort. Cancer Epidemiol Biomarkers Prev. 2019. 28(8):1300–1307.

28. Yuan B, Schafferer S, Tang Q, Scheffler M, Nees J, Heil J, Schott S. et al. A plasma metabolite panel as biomarkers for early primary breast cancer detection. Int J Cancer. 2019. 144(11):2833–2842.

29. Plewa S, Horała A, Dereziński P, Nowak-Markwitz E, Matysiak J, Kokot ZJ. Wide spectrum targeted metabolomics identifies potential ovarian cancer biomarkers. Life Sci. 2019. 222:235–244.

30. Pichler R, Fritz J, Heidegger I, Steiner E, Culig Z, Klocker H, Fuchs D. Predictive and prognostic role of serum neopterin and tryptophan breakdown in prostate cancer. Cancer Sci. 2017. 108(4):663–670.

31. Assi N, Thomas DC, Leitzmann M, Stepien M, Chajès V, Philip T. et al. Are Metabolic Signatures Mediating the Relationship between Lifestyle Factors and Hepatocellular Carcinoma Risk? Results from a Nested Case-Control Study in EPIC. Cancer Epidemiol Biomarkers Prev. 2018. 27(5):531–540.

32. Rezig L, Servadio A, Torregrossa L, Miccoli P, Basolo F, Shintu L. et al. Diagnosis of post-surgical fine-needle aspiration biopsies of thyroid lesions with indeterminate cytology using HRMAS NMR-based metabolomics. Metabolomics. 2018. 14(10):141.

33. Eniu DT, Romanciuc F, Moraru C, Goidescu I, Eniu D, Staicu A, Rachieriu C, Buiga R, Socaciu C. The decrease of some serum free amino acids can predict breast cancer diagnosis and progression. Scand J Clin Lab Invest. 2019. 79(1-2):17–24.

34. Li J, Li J, Wang H, Qi LW, Zhu Y, Lai M. Tyrosine and Glutamine-Leucine Are Metabolic Markers of Early-Stage Colorectal Cancers. Gastroenterology. 2019. 157(1):257–259.

35. Lee W, Um J, Hwang B, Lee YC, Chung BC, Hong J. Assessing the progression of gastric cancer via profiling of histamine, histidine, and bile acids in gastric juice using LC-MS/MS. J Steroid Biochem Mol Biol. 2019. 197:105539.

36. Onesti CE, Boemer F, Josse C, Leduc S, Bours V, Jerusalem G. Tryptophan catabolism increases in breast cancer patients compared to healthy controls without affecting the cancer outcome or response to chemotherapy. J Transl Med. 2019. 17(1):239.

37. Sharma NK. Ajay Kumar, Asawari Waghmode. 2019. Design of vertical tube electrophoretic system and method to fractionate small molecular weight compounds using polyacrylamide gel matrix. Date of Publication: 01/03/2019. (Patent Application Number no: 201921000760). Publication Type. INA, The patent official Journal No-19/2018, Page no-9035. Published.

38. Liigand J, Laaniste A, Kruve A. pH Effects on Electrospray Ionization Efficiency. J Am Soc Mass Spectrom. 2017. 28(3):461–469.

39. Li XL, Li G, Jiang YZ, Kang D, Jin CH, Shi Q, Jin T. et al. Human nails metabolite analysis: A rapid and simple method for quantification of uric acid in human fingernail by high-performance liquid chromatography with UV-detection. J Chromatogr B Analyt Technol Biomed Life Sci. 2015. 1002:394–8.

40. Giovanoulis G, Alves A, Papadopoulou E, Cousins AP, Schütze A, Koch HM, Haug LS. et al. Evaluation of exposure to phthalate esters and DINCH in urine and nails from a Norwegian study population. Environ Res. 2016. 151:80–90.

41. Krumbiegel F, Hastedt M, Westendorf L, Niebel A, Methling M, Parr MK, Tsokos M. The use of nails as an alternative matrix for the long-term detection of previous drug intake: validation of sensitive UHPLC-MS/MS methods for the quantification of 76 substances and comparison of analytical results for drugs in nail and hair samples. Forensic Sci Med Pathol. 2016. 12(4):416–434.

42. Alves A, Covaci A, Voorspoels S. Method development for assessing the human exposure to organophosphate flame retardants in hair and nails. Chemosphere. 2017. 168:692–698.

43. Cappelle D, Neels H, De Keukeleire S, Fransen E, Dom G, Vermassen A, Covaci A, Crunelle CL, van Nuijs ALN. Ethyl glucuronide in keratinous matrices as biomarker of alcohol use: A correlation study between hair and nails. Forensic Sci Int. 2017. 279:187–191.

44. Craig J, Ceballos DM, Fruh V, Petropoulos ZE, Allen JG, Calafat AM. et al. Exposure of nail salon workers to phthalates, di(2-ethylhexyl) terephthalate, and organophosphate esters: A pilot study. Environ Sci Technol. 2019. doi: 10.1021/acs.est.9b02474.

45. Laemmli UK. Nature. 1970. 227:680–685.

46. Mitruka M, Gore CR, Kumar A, Sarode SC, Sharma NK. Undetectable free aromatic amino acids in nails of breast carcinoma: Biomarkers discovery by a novel metabolite purification VTGE system. 2019. BioRxiv. BIORXIV/2019/876441.

